# Thoracic discopathy and thoracic-chest related pain: a scoping review

**DOI:** 10.1101/2024.03.20.24304370

**Authors:** Ferrulli Lucia, Zaninetti Martina, Margelli Michele, Maselli Filippo, Giovanni Galeoto

## Abstract

**Background:** Thoracic discopathy refers to the degeneration or herniation of one or more discs in the thoracic spine, which can cause thoracic-chest related pain. Thoracic-chest related pain can be challenging to diagnose and treat, as it can have various causes, including musculoskeletal, neurological, and visceral.

**Objectives:** This scoping review aims to provide an overview of the current scientific literature on the thoracic discopathy and the thoracic-chest related pain, by examining the available scientific research, as well as to identify any existing gaps in knowledge.

**Eligibility criteria:** The databases of Medline, Cinahl, Cochrane, Prospero were searched using key terms: “thoracic”, “chest”, “dorsal”, “disc”, “hernia”, “radiculopathy”, “myelopathy”, and “pain”.

We also have searched for Grey literature on clinicaltrial.gov and google scholar.

**Inclusion criteria:** adult population (over 18 years old); thoracic discopathy or thoracic-chest related pain; Italian or English language; no context, geographical or temporal limits. Exclusion criteria: unspecified pathologies and symptoms (as non-specific back pain, low back pain) and specific pathologies without any interest of thoracic disc (as spinal synovial cysts, spinal arachnoid webs, lung herniation ecc).

## INTRODUCTION

Thoracic discopathy, a degenerative condition affecting the thoracic spine, can cause thoracic-chest related pain, which can be debilitating and impact the quality of life of those affected. Despite this, there is a lack of comprehensive reviews of the literature on these conditions.

A scoping review is an appropriate method to address this gap in the literature as it allows for a broad exploration of the research available on a topic, including both quantitative and qualitative studies, and can help identify research gaps and areas for future research. The scoping review will aim to identify the existing evidence on the causes, symptoms, diagnosis, and treatment of thoracic discopathy. This information can inform clinicians, researchers, and patients on the current state of knowledge and potentially guide the development of future research and clinical practice guidelines.

### Research question

Identifying the current state of the literature on thoracic pain and thoracic-chest related pain associated with thoracic discopathy.

### Specific objectives

Describing the available evidence on the symptoms, causes, diagnosis, and treatment of thoracic discopathy.

### Patient

female and male over 18 years old who have thoracic discopathy.

### Concept

thoracic discopathy with thoracic-chest related pain associated to herniation, bulging, protrusion, radiculopathy or myelopathy

### Context

no limits

## METHODS

Protocol and Registration: https://www.medrxiv.org

### Eligibility criteria

Inclusion criteria: any type of published work (no limits based on study design). No limits on date or geographic location were imposed. Articles in English and Italian, or with at least an abstract in English will be considered. All studies that do not meet the above-mentioned PCC were excluded. The rationale for this selection criteria aligns with our research question.

The search was conducted on the following databases: Medline, CINAHL, Cochrane. In addition, research protocols were searched on PROSPERO.

We also have searched for Grey literature on open gray.eu and Google Scholar.

### Information sources

The search was conducted on the following databases: Medline, CINAHL, Cochrane. In addition, research protocols were searched on PROSPERO and clinicaltrial.gov.

We also have searched for Grey literature on open gray.eu and Google Scholar.

### Search

As recommended in all JBI types of reviews and PRISMA-S, a three-step search strategy is to be utilized.

- The first step is an initial limited search of an appropriate online database relevant to the topic (Pubmed). This initial search is then followed by an analysis of the text words contained in the title and abstract of retrieved papers, and of the index terms used to describe the articles.
- A second search using all identified keywords and index terms should then be undertaken across all included databases.
- Thirdly, the reference list of identified reports and articles should be searched for additional sources.
- The search strategies was peer-reviewed by an experienced librarian and further refined through team discussion
- No search limitations and filters applied (language and time)
- reviewers’ intent to contact authors of primary sources or reviews for further information
- complete search strategy for at least one major database should be included as an appendix to the protocol;
- search strategy, will be adapted for use in other database.

### Selection of sources of evidence

Selection process is based on title and abstract by two independently reviewers.

Disagreements on study selection and data extraction will be discussed with other one reviewer if needed.

Selection is performed based on inclusion criteria pre-specified.

The software used for the management of the results is Rayyan

Separate appendices for details of included and a brief mention of the excluded sources are given.

For excluded studies, reasons should be stated on why they were excluded.

There should be a narrative description of the process accompanied by a flowchart of review process (from the PRISMA-ScR statement).

Pilot testing of source selectors

- Random sample of 25 titles/abstracts is selected.
- The entire team screens these using the eligibility criteria and definitions/elaboration document.
- Team meets to discuss discrepancies and make modifications to the eligibility criteria and definitions/elaboration document.
- Team only starts screening when 75% (or greater) agreement is achieved.

When screening started it will be used a Microsoft Excel file to manage and catalogue the articles resulting from the search.

### Data charting process

Team trial the extraction form on two or three sources to ensure all relevant results are extracted, by at least two members of the review team.

Pilot step: the extraction form will be tested on two or three sources to ensure all relevant results are extracted, by two blinding members of the review team.

Inconsistencies were resolved by a third reviewer.

This form will be reviewed by the research team and pre-tested by all reviewers before implementation to ensure that the form captures the information accurately.

Modifications will be detailed in the full scoping review.

Key informations will described in a charting table with description of:

Authors

Year of publication

Country of publication

Study design

Aims of the article

Characteristics of the population (age, gender and social variables)

Definitions of “thoracic discopathy” and “thoracic chest-related pain”

Thoracic vertebral level with discopathy

Type of discopathy (herniation, bulging, protrusion, radiculopathy, myelopathy)

Type of diagnoses (clinical evaluations, imaging techniques used)

Sign and symptoms

Localization of the symptoms

Quality of the symptoms

Intensity of the symptoms

Type of referral

Type of treatment proposed: conservative or surgical

Type of conservative treatment: methods, dosage

## Data Availability

All data produced are available online at the following databases: medline, cinahl, cochrane, prospero, clinicaltrial.gov, google scholar.

## Appendix 1

Search conducted on 11st of March 2024

| DATABASE | RESULTS |
| --- | --- |
| MEDLINE<br>((thoracic OR chest OR dorsal) AND (disc OR discopathy OR Hernia OR radiculopathy OR myelopathy OR mielopathy) AND (pain)) | 3609 |
| CINAHL<br>AB ( (thoracic OR chest OR dorsal) ) AND AB ( (disc OR discopathy OR hernia OR radiculopathy OR myelopathy OR mielopathy) ) AND AB (pain) | 169 |
| COCHRANE<br>((thoracic OR chest OR dorsal) ) AND ( (disc OR discopathy OR hernia OR radiculopathy OR myelopathy OR mielopathy) ) AND (pain)) | 10 (c. reviews)<br>459 (trials) |
| PROSPERO<br>( thoracic OR chest OR dorsal ) AND ( disc OR discopathy OR hernia OR radiculopathy OR myelopathy OR mielopathy ) AND (pain) | 134 |

